# Long-term follow-up of the public health impacts and co-benefits of an urban greenway intervention: A 15-year natural experiment evaluation

**DOI:** 10.64898/2026.04.08.26350381

**Authors:** Duyen Nguyen, Christopher Tate, Selin Akaraci, Ruoyu Wang, Frank Kee, Shay Mullineaux, Ciaran O’Neill, Claire Cleland, Brendan Murtagh, Geraint Ellis, Dominic Bryan, Alberto Longo, Leandro Garcia, Mike Clarke, Ruth F. Hunter

**Author notes:** **Correspondence to:** Ruth Hunter and Duyen Nguyen, Centre for Public Health, Queen’s University Belfast, Belfast, United Kingdom (BT12 6BA). **Authors information:** Christopher Tate.

## Abstract

**Background:** Evidence on the long-term impact of urban green and blue spaces (UGBS) interventions remains limited. This study is a 15-year evaluation of an urban greenway development in Belfast (United Kingdom), assessing the potential effects of this UGBS intervention on physical activity (PA), mental wellbeing and co-benefits.

**Methods:** Using quasi-experimental design, a repeated cross-sectional survey was conducted in 2010 (baseline), 2017 (post-opening) and 2023 (long-term follow-up) with about 1,200 adults participated each wave. Outcomes included PA, mental wellbeing, general health, quality of life, social capital and environmental perception. Multilevel mixed-effect regressions were performed to examine within-group changes at long-term follow-up. Difference-in-differences analysis investigated the between-group changes that might be attributed to the greenway. Additional comparative analyses included distance-decay analysis and comparison with population trends in Northern Ireland.

**Results:** At six years after completion, the greenway intervention appears to buffer a decline in duration of PA – mainly from moderate-intensity activity (decline lower by 118.6 min/week, 95%CI: 3.9-232.2) but with no significant impact on the proportion of the population meeting the recommended PA level. The intervention is associated with a smaller decline in self-rated health (4.98 units; 95%CI: 0.62-9.34) relative to control group. Intervention association with mental wellbeing was positive but not significant (p=0.30). The greenway also showed positive effects on social capital and environmental perceptions, with impacts most evident in improving safety and trust in the local area.

**Conclusion:** This study provides evidence to support the public health impact of UGBS and its long-term health and social benefits.

## 1. INTRODUCTION

Evidence demonstrates that urban green and blue spaces (UGBS) have the potential to contribute significantly to public health, and generate a range of other co-benefits impacting social, environmental, and economic outcomes, as well as mitigating against worsening health inequalities [1-3]. Many policy frameworks have identified UGBS as a key component to promote urban regeneration and create social spaces, improving the health and mental wellbeing of the general population [4-6]. Research also demonstrates that high-quality UGBS (e.g., characterised by aesthetics, safety, and amenities) are associated with better health outcomes [7, 8]. However, these benefits are not experienced equally by all, given spatial and social inequities in the distribution of UGBSs [9, 10].

To date, there have been few longitudinal and interventional studies evaluating the effects of UGBS on health and other co-benefits [3, 11]. These studies have been hampered by short follow-up periods (i.e., 1-2 years) and have failed to adequately assess the potential impact of UGBS on overall health, non-communicable diseases (NCDs) and health inequalities in the long-term [3, 11]. As such, there is a clear need for studies of the effects of UGBS interventions which employ longitudinal designs with sufficient long-term follow-up.

For the present study, we conducted a natural experiment of an UGBS intervention, the Connswater Community Greenway (CCG) in Belfast, United Kingdom (UK) [12]. Using a quasi-experimental design, this study aimed to assess the long-term impact of an UGBS intervention 6 years after its opening to the public, and 15 years after the baseline. The primary objective was to investigate the effects of the intervention on physical activity (PA), mental wellbeing, quality of life, social capital, and environmental perceptions.

## 2. METHOD

### 2.1 Study context

The CCG was developed in east Belfast, an area with higher levels of socio-economic deprivation, older age profile, elevated death rates due to cancer, respiratory disease and circulatory diseases, and higher hospital admission rates for self-harm than the average of Northern Ireland [13-15]. With £40 million investment, the project included 9 km of linear greenway, 5 km of remediated watercourses, 16 km of footpaths and cycleway, 13 hectares of park space, and other infrastructure as hubs for education, tourism and heritage trails [14]. Community engagement work started in 2011 with the construction work commencing in 2013 and concluding in April 2017.

### 2.2 Study design

The present study examines the effects of the CCG on PA and other health outcomes using a quasi-experimental before-and-after design with repeated cross-sectional surveys to assess population-level changes. Data were collected in three waves: Wave 1 in 2010/2011 before the community engagement activities and construction began; Wave 2 in 2017/2018 covered almost a year after the completion of the CCG; and Wave 3 in 2023/2024 (6 years after CCG opened).

In each wave, a random sample of approximately 1200 respondents aged ≥ 16 years participated in data collection, representing approximately 1.2% of the population in 29 electoral wards of east Belfast. In this sampling frame, the intervention area covers 22 wards whose geographical centroids are within a 1-mile radius of the CCG (approximate 15-minute walk to access the CCG, a commonly used proxy measure in walkability literature for “accessible” [11]). Seven neighbour wards located beyond this radius served as the control group.

The survey employed a representative household sample to ensure comparability across the three waves and with other Northern Ireland surveys, providing a context for any changes observed in the population. Survey weighting was applied to account for the population size in each ward and sex, and age structure, and seasonality variation. The response rate was approximately 51% across the three waves, similar to the average achieved in the Health Survey Northern Ireland (HSNI) [16]. More details about the survey methods are in the published protocol [11].

### 2.3 Outcome measures & covariates

The primary outcome was change in PA, assessed using the Global Physical Activity Questionnaire (GPAQ) [17] as: 1) percentage of people meeting the recommended 150 minutes of moderate-intensity PA per week; and 2) total time of PA (converted to equivalent of moderate-intensity PA minutes/week). The pattern of PA was also inspected in specific domains (types and intensities) including: 1) time spent in vigorous-intensity PA; 2) time spent in moderate-intensity PA (excluding time for active travel); 3) time spent in active travel (e.g., walking/cycling); and 4) percentage of people using active travel modes such as walking/cycling during a week.

Secondary outcomes included health and wellbeing, social capital, and environmental perceptions. Health and wellbeing outcomes measured: 1) mental wellbeing using the Warwick Edinburgh Mental Wellbeing Scale (WEMWBS) [18]; 2) general health using the Short Form-8 Health Survey (SF-8) [19]; and 3) quality of life using the EQ-5D tool providing overall self-rated health (EQ-VAS) and health-related quality of life (EQ-5D-3L) [20].

Social capital and environmental perceptions were measured using the instrument from the UK General Household Survey [21, 22]. Social capital was assessed with respect to local area trust and social network. Perceptions of the environment focused on: 1) perceived attractiveness of the area; 2) traffic levels and congestion; 3) availability of local amenities; and 4) safety in the neighbourhood.

All analyses were adjusted for the following covariates: gender; age; marital status; education; employment status; accommodation type; car ownership; long-term illness; and household income. Additional details about the study outcomes and covariates are provided in Appendix 1.

### 2.4 Comparators

To reduce risk of bias, as well as having a control group sampled from seven wards adjacent to the study site, the study employed two other comparators for analysis [23]. The first comparator used the walking distance from the participant’s household to the nearest CCG access point (0-400m, 400–800m, 800–1200m, and >1200m) as a proxy measure of intervention exposure to perform a distance-decay analysis. Second, trends in the intervention group were compared with population trends in Northern Ireland using data from the HSNI [16] in terms of PA and mental wellbeing (WEMWBS). However, we acknowledge that HSNI used the International Physical Activity Questionnaire and measurement was only available in a few years, so this comparison needs cautious interpretation.

### 2.5 Statistical analysis

The primary statistical analysis comprised three parts: 1) multilevel mixed-effects regression analysis to examine the within-group changes in outcomes over time; 2) difference-in-differences (DID) models to estimate the between-group changes that were associated with the impact of the CCG; and 3) distance-decay analysis performed separately for the intervention group as a sensitivity analysis.

The multilevel regression analysis examined the long-term change in each outcome between waves 1 and 3. This allowed us to investigate the direction and magnitude of within-group changes in the intervention group and control group separately. All multilevel regression models included a random intercept at the super output area level to account for clustering of individuals within areas and were adjusted for the covariates listed above. For binary outcomes, the mean difference was assessed using odds ratios (OR) obtained from a multilevel logistic regression model. For other outcomes, mean differences were reported using the coefficients obtained from multilevel mixed-effect linear regressions.

The DID approach used two models: 1) unmatched-DID and 2) propensity score matching DID (PSM-DID) to estimate the between-group changes that could be attributed to a CCG effect. The first is a conventional DID in the form of a linear regression model with a DID interaction term included, representing the effect of the CCG. In the PSM-DID model, the matching process for repeated cross-sectional data was performed using pre-intervention covariates to balance the intervention and control group at baseline [24]. These weights were then applied to both pre- and post-intervention samples using a kernel function with a bandwidth of 0.6 to ensure comparability of the two groups over time.

The distance-decay analysis was conducted in the intervention group only by stratifying participants into four groups by distance to the CCG: <400m, 400-800m, 800-1200m, and >1200m radius. This grouping corresponds to an approximate 5-, 10-, and 15-minute walk from the participant’s home to the nearest CCG access point – representing different levels of exposure to the intervention that have been used in existing literature on environmental interventions [25]. Multilevel regression models were performed in which distance-decay was tested using distance-band × wave interaction terms (joint Wald test).

All results were weighted to reflect the composition of the whole population by ward-level population size, gender, age and seasonality. All analyses were conducted in STATA version 17.0 with a significance level of 5%.

## 3. RESULTS

### 3.1 General characteristics

In the long-term follow-up sample, 47.9% of the intervention group and 48.7% of the control group were male, with corresponding mean ages of 47.41 years and 49.03 years, respectively. Both groups had a similar gender and age profile across all three waves (see **Table 1**). There was a slight reduction over time in the proportion of participants from more deprived areas and lower income quintiles in the intervention group, but it remained overall more deprived than the control group.

**Table 1:** Participant characteristics across three waves of household survey*.

| Year<br>*n* = | Intervention |  |  | Control |  |  |
| --- | --- | --- | --- | --- | --- | --- |
|  | Wave 1<br>1037 | Wave 2<br>968 | Wave 3<br>944 | Wave 1<br>168 | Wave 2<br>246 | Wave 3<br>258 |
| **Gender** |  |  |  |  |  |  |
| Male | 47.7% | 48.2% | 47.9% | 44.1% | 45.1% | 48.7% |
| Female | 52.3% | 51.8% | 52.1% | 55.9% | 54.9% | 51.3% |
| **Age (mean, SD)** | 46.1 (18.6) | 47.7 (18.1) | 47.4 (17.5) | 50.6 (19.1) | 49.3 (18.2) | 49.0 (17.3) |
| **Marital status** |  |  |  |  |  |  |
| Single | 31.6% | 35.3% | 11.5% | 23.3% | 25.6% | 8.0% |
| Married/Cohabited | 48.7% | 44.8% | 39.0% | 56.6% | 57.2% | 53.2% |
| Other | 19.7% | 19.9% | 49.5% | 20.1% | 17.2% | 38.8% |
| **Education <sup>a</sup>** |  |  |  |  |  |  |
| High education | 41.3% | 36.6% | 33.8% | 40.5% | 36.2% | 33.0% |
| Low education | 58.7% | 63.4% | 66.2% | 59.5% | 63.8% | 67.0% |
| **Employment** |  |  |  |  |  |  |
| Employed | 57.0% | 56.8% | 62.6% | 51.6% | 54.9% | 63.8% |
| Unemployed/other | 43.0% | 43.2% | 37.4% | 48.4% | 45.1% | 36.2% |
| **Accommodation types** |  |  |  |  |  |  |
| Owned | 30.5% | 26.8% | 31.3% | 37.2% | 33.6% | 35.4% |
| Mortgaged | 34.4% | 28.6% | 34.3% | 41.1% | 39.0% | 40.8% |
| Rented/other | 35.2% | 44.6% | 34.5% | 21.8% | 27.4% | 23.8% |
| **Car ownership** |  |  |  |  |  |  |
| None | 26.2% | 28.8% | 20.3% | 20.1% | 19.1% | 12.6% |
| 1 | 44.5% | 44.2% | 47.2% | 46.7% | 46.0% | 43.7% |
| 2 or more | 29.3% | 27.0% | 32.5% | 33.2% | 34.9% | 43.7% |
| **Long-term illness** |  |  |  |  |  |  |
| No | 71.5% | 71.5% | 71.1% | 70.8% | 78.4% | 73.9% |
| Yes | 28.5% | 28.5% | 28.9% | 29.2% | 21.6% | 26.1% |
| **Household income** |  |  |  |  |  |  |
| 1 (poorest) | 34.2% | 22.6% | 17.5% | 33.6% | 24.4% | 15.2% |
| 2 | 10.4% | 12.5% | 19.0% | 8.5% | 10.7% | 15.1% |
| 3 | 26.1% | 34.6% | 14.6% | 32.9% | 29.4% | 13.7% |
| 4 | 11.2% | 13.1% | 15.7% | 11.2% | 14.1% | 17.6% |
| 5 (richest) | 18.2% | 17.2% | 33.3% | 13.9% | 21.6% | 38.4% |
| **Area-based deprivation <sup>b</sup>** |  |  |  |  |  |  |
| 1 (most deprived) | 27.3% | 23.9% | 21.8% | 0.0% | 0.0% | 0.0% |
| 2 | 17.6% | 18.0% | 13.5% | 40.5% | 40.4% | 36.7% |
| 3 | 20.4% | 20.5% | 12.0% | 0.9% | 1.0% | 0.0% |
| 4 | 18.3% | 21.8% | 20.1% | 27.6% | 37.7% | 0.0% |
| 5 (least deprived) | 16.3% | 15.8% | 32.6% | 31.0% | 20.9% | 63.3% |
| **Distance to CCG <sup>c</sup>** |  |  |  |  |  |  |
| <400m | 25.3% | 21.5% | 20.9% | N/A | N/A | N/A |
| 400 – 800m | 26.9% | 24.2% | 24.6% |  |  |  |
| >800 – 1200m | 22.2% | 20.2% | 19.5% |  |  |  |
| >1200m | 25.6% | 34.1% | 35.0% |  |  |  |

\* Estimated with survey weights to account for population size, gender, age and seasonality.
a- Education (high: obtained a degree or A-level vs. low level: apprenticeship/ GCSE/ none);
b- Area-based deprivation was obtained at the super output area (SOA) level of Northern Ireland Multiple Deprivation Measure 2010, hence, the control group sampled from 7 wards did not cover all deprivation quintiles.
c - Distance to CCG was estimated only for the intervention group.

### 3.2 Effect of the CCG

Results of the multilevel mixed-effect regressions and DID analyses are presented in **Table 2**. The multilevel models show the within-group change in the intervention and control groups relative to baseline, while the DID models provide the estimated between-group changes associated with the CCG. Detailed findings are discussed separately below for PA, health and wellbeing, and social capital and environmental perceptions.

**Table 2:** Change of study outcomes at long-term follow-up as compared to baseline.

| Wave 3 vs Wave 1 | Within-group change: Multilevel mixed-effect models |  |  |  |  |  | Between-group change: DID models |  |  |  |  |  |
| --- | --- | --- | --- | --- | --- | --- | --- | --- | --- | --- | --- | --- |
|  | INTERVENTION |  |  | CONTROL |  |  | Unmatched DID |  |  | PSM-DID |  |  |
|  | Mean change* | 95%CI | P | Mean change* | 95%CI | P | Est. DID** | 95%CI | p | Est. DID** | 95%CI | p |
| **Physical activities** |  |  |  |  |  |  |  |  |  |  |  |  |
| % Meet the PA target | 0.96 | (0.73, 1.27) | 0.79 | 1.15 | (0.75, 1.76) | 0.53 | -0.03 | (-0.13, 0.07) | 0.53 | -0.04 | (-0.12, 0.03) | 0.27 |
| % Had active travel <sup>a</sup> | **1.65** | **(1.33, 2.06)** | **<0.001** | **1.84** | **(1.06, 3.22)** | **0.03** | -0.03 | (-0.15, 0.08) | 0.55 | -0.05 | (-0.12, 0.03) | 0.25 |
| Total time for PA <sup>b</sup> | **-140.5** | **(-268.7, -12.3)** | **0.03** | **-210.4** | **(-421.8, 1.1)** | **0.05** | 108.9 | (-185.9, 403.7) | 0.47 | 101.2 | (-89.0, 291.4) | 0.30 |
| Time of vigorous activities | -18.5 | (-57.7, 20.8) | 0.36 | 0.73 | (-107.2, 108.6) | 0.99 | -21.4 | (-118.6, 75.7) | 0.67 | -7.9 | (-70.0, 54.2) | 0.80 |
| Time of moderate activities <sup>c</sup> | **-103.8** | **(-171.8, -35.7)** | **0.003** | **-244.2** | **(-435.5, -53.0)** | **0.012** | 154.8 | (-46.7, 356.3) | 0.13 | **118.0** | **(3.9, 232.2)** | **0.043** |
| Time for active travel | -2.3 | (-39.9, 35.3) | 0.91 | 31.4 | (-67.2, 130.1) | 0.53 | -3.0 | (-94.4, 88.4) | 0.95 | -1.0 | (-52.4, 50.3) | 0.97 |
| **Health and wellbeing** |  |  |  |  |  |  |  |  |  |  |  |  |
| WEMWBS score <sup>d</sup> | -0.92 | (-1.88, 0.05) | 0.06 | -1.76 | (-3.81, 0.29) | 0.09 | 0.69 | (-1.32, 2.69) | 0.50 | 0.80 | (-0.72, 2.32) | 0.30 |
| SF-8: Physical component | 0.71 | (-0.35, 1.77) | 0.19 | 0.57 | (-1.28, 2.42) | 0.55 | 0.09 | (-1.89, 2.07) | 0.93 | 0.14 | (-1.74, 2.02) | 0.89 |
| SF-8: Mental component | **1.80** | **(0.79, 2.81)** | **<0.001** | 2.28 | (-0.61, 5.16) | 0.12 | -0.37 | (-2.83, 2.09) | 0.77 | -0.20 | (-2.14, 1.74) | 0.84 |
| EQ-VAS <sup>e</sup> | **-10.41** | **(-16.24, -4.59)** | **<0.001** | **-14.74** | **(-26.6, -2.8)** | **0.015** | 0.02 | (-0.02, 0.05) | 0.41 | **4.98** | **(0.62, 9.34)** | **0.025** |
| EQ5D3L | **-0.06** | **(-0.08, -0.03)** | **<0.001** | **-0.09** | **(-0.15, -0.03)** | **0.005** | 0.04 | (-0.01, 0.08) | 0.13 | 0.03 | (-0.02, 0.08) | 0.22 |
| **Social capital** |  |  |  |  |  |  |  |  |  |  |  |  |
| Local area trust <sup>f</sup> | **-1.82** | **(-2.01, -1.64)** | **<0.001** | **-2.19** | **(-2.42, -1.96)** | **<0.001** | **0.37** | **(0.25, 0.50)** | **<0.001** | **0.30** | **(0.20, 0.39)** | **<0.001** |
| Social network <sup>g</sup> | **-1.03** | **(-1.10, -0.97)** | **<0.001** | **-0.87** | **(-0.98, -0.77)** | **<0.001** | -0.11 | (-0.24, 0.02) | 0.11 | **-0.13** | **(-0.23, -0.04)** | **0.007** |
| **Environmental perceptions** |  |  |  |  |  |  |  |  |  |  |  |  |
| Attractive <sup>h</sup> | **0.29** | **(0.21, 0.38)** | **<0.001** | **0.21** | **(0.04, 0.39)** | **0.015** | 0.12 | (-0.03, 0.28) | 0.12 | 0.07 | (-0.05, 0.19) | 0.25 |
| Traffic <sup>h</sup> | **0.28** | **(0.20, 0.36)** | **<0.001** | **0.24** | **(0.04, 0.45)** | **0.022** | 0.09 | (-0.08, 0.27) | 0.31 | 0.10 | (-0.02, 0.23) | 0.10 |
| Amenities <sup>h</sup> | **-0.36** | **(-0.43, -0.30)** | **<0.001** | **-0.30** | **(-0.46, -0.15)** | **<0.001** | 0.02 | (-0.13, 0.17) | 0.77 | 0.03 | (-0.07, 0.13) | 0.54 |
| Safety <sup>h</sup> | **-1.39** | **(-1.57, -1.20)** | **<0.001** | **-1.61** | **(-1.98, -1.24)** | **<0.001** | **0.25** | **(0.08, 0.43)** | **0.005** | **0.25** | **(0.12, 0.38)** | **<0.001** |

PA=physical activity, EQ5D3L = European Quality of life 5-dimension 3-level, EQ-VAS = Visual analogue scale, DID=difference in difference, PSM-DID: DID with propensity score matching.
\* Mean change: % meeting the PA target and % had active travel reported OR from the multilevel logistic model. For other outcomes, coefficients from multilevel linear regressions were reported.
\*\* Estimated DID: For % meeting the PA target and % had active travel, DID reported coefficients of linear probability regressions. For other outcomes, DID was obtained from linear models.
a – having walking/ cycling during a typical week; b - equivalent of minutes of moderate-intensity PA per week; c - Moderate activities at work + recreational activities, not including active travel; d - WEMWBS scale from 14–70; e - EQ-VAS scale from 0–100; f - 1=very big problem to 4=not a problem at all; g - 1=never to 5=most days; h - 1=strongly disagree to 5=strongly agree.

#### 3.2.1. Physical activity

The study found a significant association between the presence of CCG and a reduction in the decline in time spent in moderate-intensity PA, which led to a positive shift in total time of PA. However, we did not find a net impact of CCG on the proportion of the population meeting the PA recommendation.

Specifically, both the intervention and control group showed significant reduction compared to baseline in time spent in moderate-intensity PA: 103.8 minutes/week (95%CI : 35.7, 171.8) and 244.2 minutes/week (95%CI: 53.0, 435.5), respectively. After adjusting for other factors, the results suggest that the CCG may have significantly mitigated the decline, reducing the decrease in time for moderate-intensity PA by 118.0 min/week (95%CI: 3.9, 232.2).

In regard to total time of PA, the multilevel model showed a significant reduction in total time for PA for both the intervention group (-140.5 min/week, 95%CI: -268.7, -12.3) and control group (-210.4 min/week, 95%CI: -421.8, 1.1). The time reduction was less pronounced in the presence of CCG (mean DID of 101.2 min/week for intervention versus control group, p=0.30), but the impact was not statistically significant (95%CI:-89.0, 291.4).

The proportion of the population meeting the recommended PA guidelines had not changed significantly at long-term follow-up, either in the intervention group (OR=0.96, 95%CI: 0.73-1.27) or the control group (OR=1.15, 95%CI: 0.75, 1.76). This finding aligns with the population trend reported by the HSNI in 2023/24, as compared to 2012/2013 or 2016/2017 (see Appendix 3) [16]. The DID analysis also confirmed that the CCG did not have a significant impact on this measure.

#### 3.2.2. Health and wellbeing

**Table 2** shows that the CCG was associated with an overall positive impact on health and wellbeing, but these potential impacts were relatively small and varied across three measurements. WEMWBS and EQ-5D-3L scores declined over time but a smaller reduction was observed in the intervention group. General health (both physical and mental components) measured using SF-8 had increased for the intervention and the control group at long-term follow up, but no significant impact attributable to the CCG was observed.

WEMWBS scores of the intervention group exhibited a non-significant reduction (-0.92 points, 95%CI: -1.88, 0.05) 6 years after completion of the CCG, which was similar to the control group (-1.76 points, 95%CI: -3.81, 0.29) – reflect the stable trend across Northern Ireland [16] (see Appendix 3). The DID analysis aligned with this finding, showing a protective effect, albeit not statistically significant, in both unmatched DID (mean DID = 0.69, p=0.50) and PSM-DID (mean DID = 0.80, p=0.30) models.

The EQ-5D tool showed significant declines in both self-rated health (EQ-VAS) and quality of life (EQ5D3L) for both the intervention and control group. For the intervention group, self-rated health declined by 10.41 (95%CI: 4.59, 16.42) and quality of life decreased by 0.06 (95%CI: 0.03, 0.08). The reduction was also significant but numerically higher for the control group, with a decline of 14.74 points (95%CI: 2.8, 26.6) in self-rated health and 0.09 points (95%CI: 0.03, 0.15) in quality of life. The matched DID models indicated a positive impact of the CCG for both self-rated health and quality of life; but the effect was statistically significant only for self-rated health (mean DID = 4.98 points, p=0.025).

Using the SF-8 tool, no significant changes were observed in either the physical or mental health components for either group, except for an increase in mental functional health in the intervention group. However, the DID analysis indicated that the CCG had no significant effect on either component of the SF-8.

#### 3.2.3. Co-benefits: social capital and environmental perceptions

The results suggest that the CCG was associated with less of a decline in trust in the local area among the respondents (see **Table 2**). This finding is positive despite the significant decrease in trust for both groups relative to baseline (intervention: -1.82 units versus control: -2.19 units, both with p<0.001). The DID model confirmed this, estimating that the CCG offset the decline by 0.30 units (p<0.001). In contrast, while social networks declined across both groups, the DID analysis indicated the decline was greater in the intervention group after completion of the CCG (mean DID -0.13, p=0.007).

The study found a generally favourable association between the CCG and all four aspects of environmental perception (see DID models in **Table 2**), but the impact reached statistical significance only for perceived safety. The DID analysis suggested that despite a substantial decline in perceived safety, the CCG offered a protective effect, mitigating the decline by 0.25 points (on a 5-point scale) (p=0.005 in unmatched DID, p<0.001 in PSM-DID model).

### 3.3 Distance-decay effects

The distance-decay effect analysis (**Table 3**) found no distance-decay effect of the CCG on most outcomes, except for local area trust and perceived safety. Specifically, compared to baseline, the reduction in perception of safety was larger as participants lived further from CCG, from a reduction of 1.20 units (0-400m) to 1.27 (400-800m and 800-1200m) and to 1.67 (>1200m) (p<0.001 for all). During the post-intervention period, for those who lived 400m, 800m, 1200m and >1200m from the CCG, the perception of safety fell by 1.35, 1.45, 1.46 and 1.92 units, respectively. A similar pattern was observed for the reduction in local area trust during the post-intervention period.

**Table 3:**
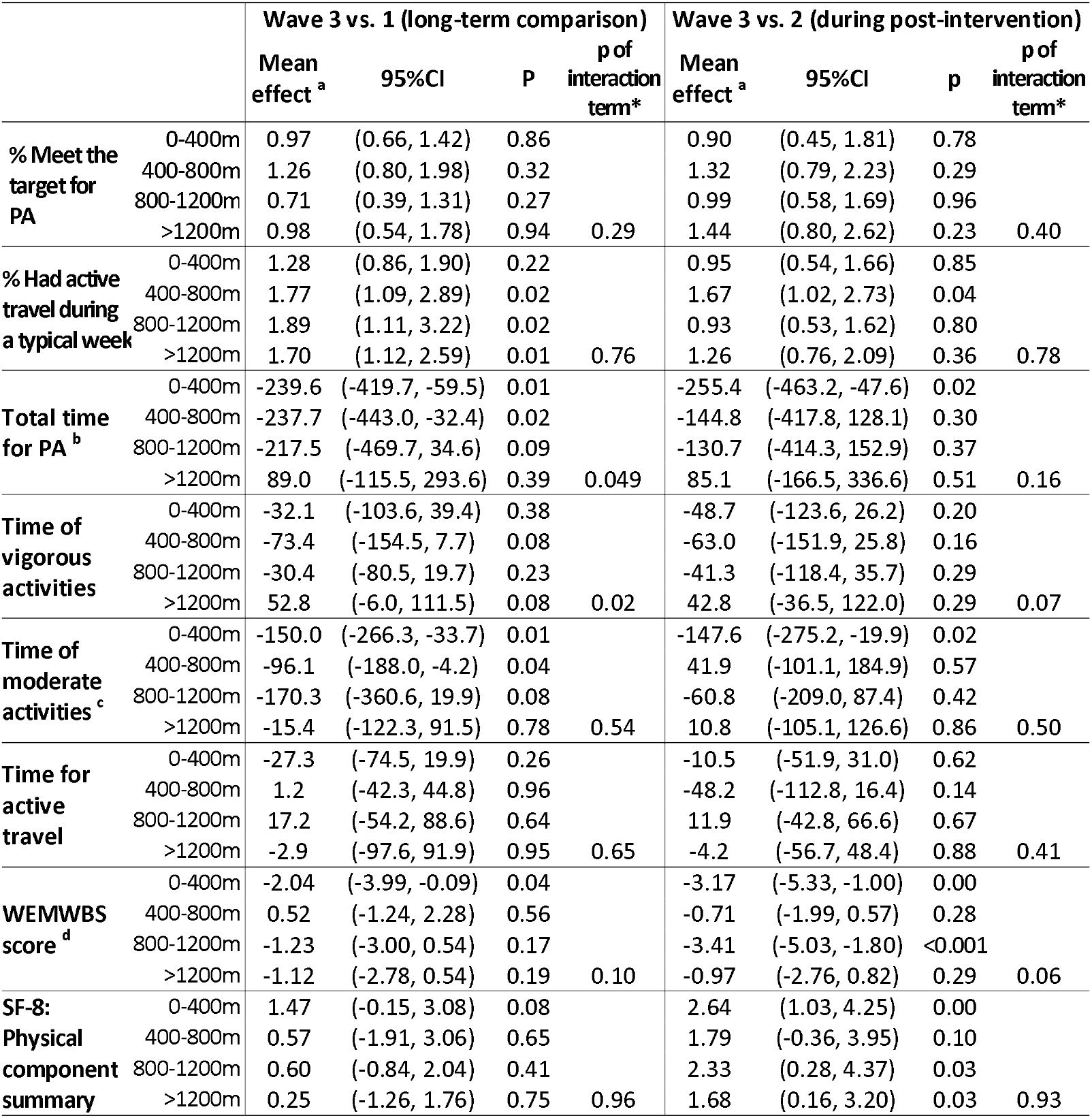
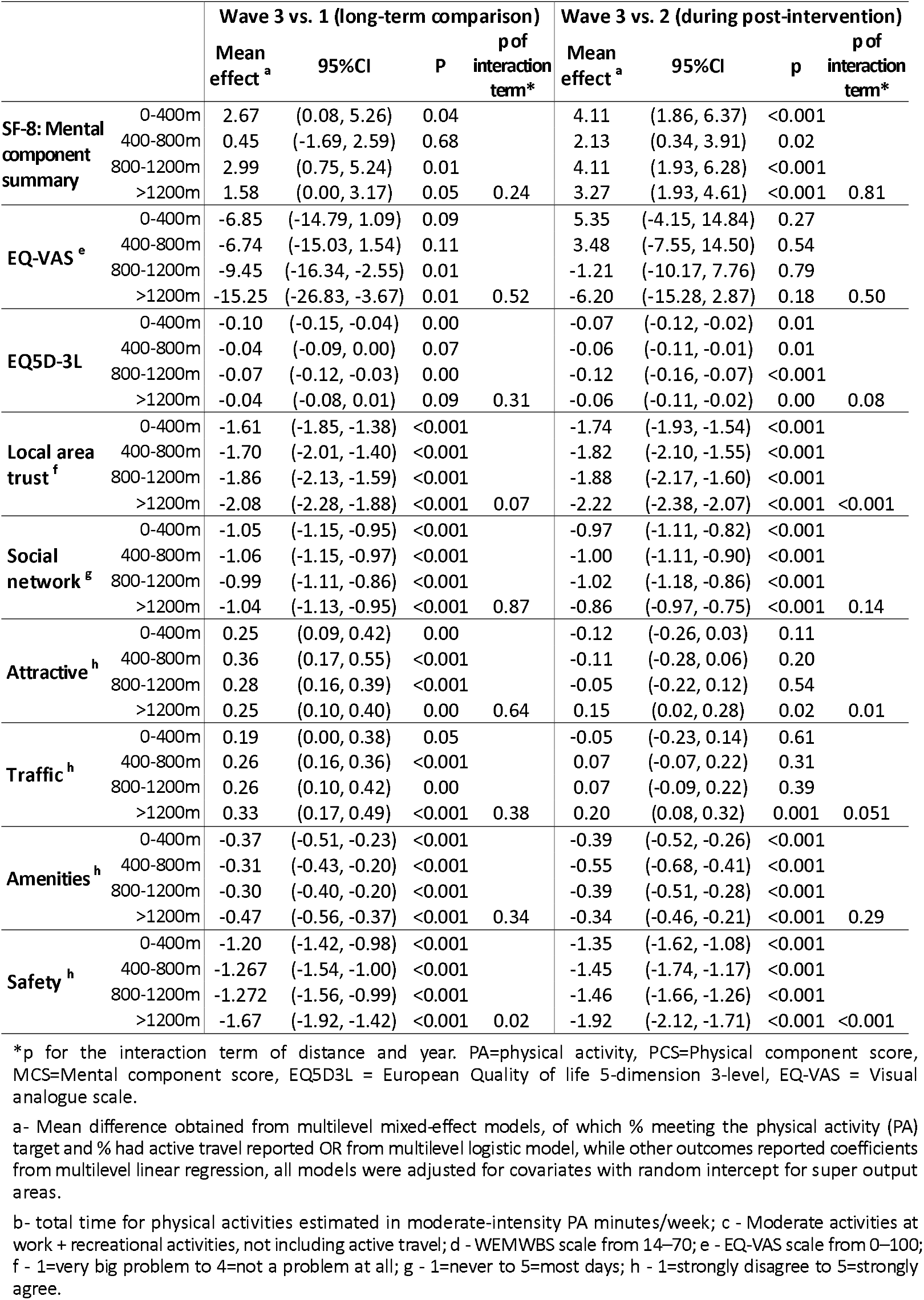
The difference in mean outcomes across three waves based upon the distance to CCG (intervention sample)

## 4. DISCUSSION

### 4.1 Physical activity

Overall, this study has shown that six years after completed, the CCG is associated with less decline in duration of PA in the intervention group relative to the control group, but there was no significant change in the proportion of population meeting the PA recommendations. This result was driven mainly by time spent in moderate-intensity PA which the CCG may have preserved approximately 118 minutes of moderate intensity PA per week (equivalent of 16.8 min/day). This is importance because PA declined overall in both the intervention and control group with people spending less time in PA even though the percentage of the population meeting PA recommendation did not change much after the intervention. Although the change was modest, the development of the CCG involved a prolonged five-year construction period during which there was restricted access to public spaces and a temporary decline in environmental quality which are likely to have contributed to the reduction in PA [26]. The study also found that PA levels were consistent over six years post-intervention, with a non-significant increase in the percentage of people meeting the PA recommendation, and a continuous increase in active travel since the immediate follow-up of the opening of the CCG in 2017 (see Appendix 4). We posit that these changes were a direct result of improved walkability, linked to opening multiple access points; better integration with active travel services; and an increase in community programmes to diversify use of the greenway [13].

### 4.2 Health and wellbeing

Evidence suggests that the CCG may have had an overall positive long-term impact on mental wellbeing and quality of life by mitigating against the worsening trends for these outcomes in Belfast and in Northern Ireland more generally. Quality of life and self-rated health declined in both the intervention and the control group over the short- and long-term, but the parallel trend comparison showed that the decline was less for the intervention group. There is no comparable UK data on quality of life (EQ-5D) within a similar timeframe; hence, external validation of this result is limited to our study setting. The (non-significant) declining pattern of WEMWBS scores in the intervention group was similar to the population trend [16] but better than the control group after controlling for the parallel trends. Together, the evidence indicates that the CCG may have had a marginal impact on wellbeing, which aligns with previous research, but emphasises the need to target deprived areas and low-income groups, especially to promote psychological restoration and reduce stress [2].

### 4.3 Social capital and environmental perceptions

For both the intervention and control groups, a gradual decline was observed in measures of social capital and residents’ perceptions of the living environment across the three waves. However, our findings suggest that the project may have contributed towards reversing some of the negative trends in social cohesion and environmental satisfaction. In the long-term follow-up, local area trust of those in the intervention group improved significantly compared to the control group, as did their perceptions of safety. This conclusion was also supported by the distance-decay analysis, indicating a dose-response relationship, which suggests that living closer to a well-designed UGBS can sustain trust and feelings of safety in the local area. The results also indicated that people who lived further from the CCG reported a larger decline in these aspects, with a clear gradient of impact aligned with their walking distance from the CCG. This is a positive result given that the CCG design and planning had focused heavily on increasing the safety and accessibility of UGBS for local residents. This result is consistent with previous research that identified how improvements in walkability, greenery, and public space design can contribute to perceived safety and social trust. This is consistent with previous research [27] but emphasises the potential to address trust as a barrier and enabler to extend and diversify use of greenways as a safe and accessible asset.

### 4.4 The need of long-term vision for UGBS interventions

Being the first study with more than 5 years follow-up of the health impacts of an UGBS intervention, this study demonstrates the potential for a long-term impact of CCG across multiple outcomes, as compared to the effects observed in the short term. This is of great importance given the decline in PA, health and wellbeing observed across the population. At 6 years after CCG completion, the short-term evaluation showed that the intervention may have had a protection effect on PA, mental wellbeing and quality of life [28]. The results also showed significant improvements in participants’ perceptions of environmental and social capital post-intervention[28], particularly of feelings of safety and social trust [29]. Additionally, a social return on investment analysis of the CCG found that for every £1 invested in the greenway, the local economy would see benefits valued between £1.34 and £1.59 through increased residential property values, reduced risk of flooding, increased tourism and reduced air pollution [30]. Together, the findings from this 15-year evaluation of the CCG emphasise the need for a long-term vision in urban planning, beyond typical four-year political cycles, to fully capture the benefits and potential of UGBS interventions.

### 4.5 Strengths and limitations

This study timeframe had a substantial time gap between each wave: seven years from the baseline to the first short-term evaluation, and fifteen years to the long-term follow-up reported here. Although our analysis adjusted for changes in sample demographic composition, the study was limited in accounting for broader societal shifts, such as evolving social norms, increasing health awareness, and changing attitudes toward PA during this period. The second limitation is the inability to control for the disruption caused by the COVID-19 pandemic between the wave 2 and wave 3 follow-up surveys.

## 5. CONCLUSION

This is the first study, to our knowledge, providing long-term follow-up on health and other outcomes beyond five years of the implementation of an UGBS intervention. The findings suggest modest yet meaningful positive effects at the population-level on PA, health and wellbeing, social capital, and environmental perceptions. While the overall benefits were moderate, they are particularly valuable in the context of worsening health and wellbeing trends that have been observed across multiple domains in the general population over the past decade. The study reinforces the importance of adopting a long-term perspective in both the design and evaluation of UGBS initiatives to inform future policy and practice.

## Data Availability

The datasets used and/or analysed during the current study are available from the corresponding author on reasonable request.

## List of abbreviations

CCG: Connswater Community Greenway
EQ-5D: EuroQol 5-dimension
EQ-VAS: EuroQol visual analogue scale
GPAQ: Global Physical Activity Questionnaire
HSNI: Health Survey Northern Ireland
MCS: Mental Component Score
PA: physical activity
PCS: Physical Component Score
SF-8: Short Form 8 Health Survey
UGBS: urban green and blue space
WEMWBS: Warwick Edinburgh Mental Wellbeing Scale

## ACKNOWLEDGEMENT

The study is conducted thanks to UK Prevention Research Partnership funded GroundsWell consortium. GroundsWell is an interdisciplinary consortium involving researchers, policy, implementers and communities. It is led by Queen’s University Belfast, University of Edinburgh and University of Liverpool in partnership with Cranfield University, University of Exeter, University of Glasgow, University of Lancaster and Liverpool John Moores University.

We would like to acknowledge our stakeholders including: Belfast, Edinburgh and Liverpool City Councils, Public Health Agencies of Scotland and Northern Ireland, Greenspace Scotland, Scottish Forestry, Edinburgh and Lothians Health Foundation, Department for Infrastructure Northern Ireland, Belfast Healthy Cities, Climate Northern Ireland, Health Data Research UK, Administrative Data Research Centre, NatureScot, Mersey Care NHS Foundation Trust, Liverpool City Region Combined Authority, Liverpool Health Partners, NHS Liverpool Clinical Commissioning Group, the Scottish Government, Edinburgh Health and Social Care Partnership, HSC Research and Development Office Northern Ireland, EastSide Partnership, Ashton Centre, Regenerus, Sustrans, Cycling UK, CHANGES, The Mersey Forest, Translink, Anaeko, AECOM, The Paul Hogarth Company and Moai Digital.

We also wish to acknowledge the co-investigators involved in the PARC Study (which collected the data on the wave 1 and wave 2 household surveys, and other elements of the protocol development): Mark A Tully, Helen McAneney, Margaret E Cupples, Michael Donnelly, George Hutchinson, Alberto Longo, Lindsay Prior, Michael Stevenson and Christopher Cardwell. The PARC study (which funded wave 1 and wave 2 household survey data collection) is supported by a grant from the National Prevention Research Initiative. The Funding Partners are (in alphabetical order): Alzheimer’s Research Trust; Alzheimer’s Society; Biotechnology and Biological Sciences Research Council; British Heart Foundation; Cancer Research UK; Chief Scientist Office, Scottish Government Health Directorate; Department of Health; Diabetes UK; Economic and Social Research Council; Engineering and Physical Sciences Research Council; Health and Social Care Research and Development Division of the Public Health Agency (HSC R&D Division); Medical Research Council; The Stroke Association; Welsh Assembly Government and World Cancer Research Fund.

In addition, the authors would like to acknowledge the partners and stakeholders involved in the study, including: EastSide Partnership; Belfast City Council; Department of Health, Social Services and Public Safety; Department for Regional Development; Department of the Environment; Department for Social Development; Belfast Health and Social Care Trust; East Belfast Community Development Agency; SportNI; Belfast Healthy Cities; Sustrans; Public Health Agency; Ordnance Survey NI and the local residents of the CCG population.

## Declaration of interests

### Author contributions

RH (principal investigator) had the original idea for the study and led its design of the study and the application for grant funding. DN led the data analysis, DN and RH wrote the first draft. RH, FK, CON, BM, GE, DB, AL, LG and MC contributed to the funding application and study design. SM and CC contributed to data management and GIS data element. CT, SA, RW provides critical intellectual inputs for the critical revisions of the paper. All authors read, contributed and approved the final manuscript.

### Ethical approval

The study was approved by the Office for Research Ethics Committees, Northern Ireland (09/NIR02/66).

### Consent for publication

Participants provided written informed consent prior to taking part in the study.

### Patient and public involvement (PPI statement)

Patients and/or the public were involved in the design, or conduct, or reporting, or dissemination plans of this research. Refer to the published protocol for further details (available at https://doi.org/10.1136/bmjopen-2024-097530).

