## Supplementary file for "Long-term follow-up of the public health impacts and co-benefits of an urban greenway intervention: A 15-year natural experiment evaluation"

### Appendix 1: Descriptions about study outcomes and covariates

#### Table S1: Description of study outcomes

| Outcomes | Description | Form |
|---|---|---|
| **Physical activities (using Global Physical Activity Questionnaire [1])** | | |
| % meeting the recommended PA target | % of people meeting the recommendation of at least 600 MET-min/ week (4 MET-min = 1 minute of moderate intensity activities) | Binary (1 Yes/ 0 No) |
| % active travel | % of people reported to had active travelling such as walking/ cycling/ etc during a week | Binary (1 Yes/ 0 No) |
| Total PA time per week | The total time spent on PA from work, recreational activities and active travelling using the guideline of GPAQ (4 MET-min = 1 min of moderate intensity PA) | Continuous (min/week of moderate-PA) |
| Total vigorous-intensity PA per week | Total time spent in vigorous-intensity PA at work and recreational activities during a typical week | Continuous (min/ week) |
| Total moderate-intensity PA per week | Total time spent in moderate-intensity PA at work and recreational activities | Continuous (min/ week) |
| Total active travel per week | Total time spent on active travel per week such as walking/ cycling | Continuous (min/ week) |
| **Health & Wellbeing** | | |
| Mental wellbeing | Measured using Warwick Edinburgh Mental Wellbeing Scale (WEMWBS) [2] | Continuous (range: 14-70) |
| General health | Using Short Form 8 (SF-8) with physical functional health measured using the Physical Component Score (PCS) and mental functional health using Mental Component Score (MCS) [3] | Continuous* |
| Quality of life: Self-rated health | Using the visual analogue scale (EQ-VAS) in the EuroQol 5D tool [4] | Continuous (1-100) |
| Quality of life | Utility score weighted using EuroQol 5D-3L tool [4]. Wave 1 and 2 used the version EQ5D-3L and wave 3 used the EQ5D5L. The mapping between two tools was used to obtain the quality of life in two measurements [5]. | Continuous* |
| **Social capital [6]** | | |
| Local area trust | Average of 8 items:<br>1) People being drunk or rowdy in public places?<br>2) Rubbish or litter lying around<br>3) Vandalism, graffiti and other deliberate damage to property or vehicles<br>4) People using or dealing drugs<br>5) People being attacked or harassed because of their religion (Catholic or Protestant)<br>6) People being attacked or harassed because of their skin colour, ethnic origin<br>7) Teenagers hanging around on the street<br>8) Problem with troublesome neighbours | 1- very big problem to 4 not a problem at all |

| Outcomes | Description | Form |
|---|---|---|
| Social networks | Average of 5 items:<br>1) Speak to relatives on the phone<br>2) Speak to friends on the phone<br>3) Speak to neighbours (face to face)<br>4) Meet with relatives who are not living with you<br>5) Meet up with friends | 1 never – 5 most days (scale reversed) |
| **Environmental perceptions [7]** | | |
| Attractive | Average of 3 items:<br>1) Pleasant to walk;<br>2) There is little green space;<br>3) The surroundings are unattractive | 1 strongly disagree - 5 strongly agree |
| Traffic | Average of 4 items:<br>1) There is a lot of traffic noise<br>2) The roads are dangerous for cyclists<br>3) There is little traffic<br>4) It is safe to cross the road | 1 strongly disagree - 5 strongly agree |
| Amenities | Average of 5 items:<br>1) There is a park within walking distance<br>2) There is convenient public transport<br>3) There are convenient routes for cycling<br>4) The nearest shops are too far to walk to<br>5) There are no convenient routes for walking | 1 strongly disagree - 5 strongly agree |
| Safety | Average of 2 items:<br>1) People are likely to be attacked<br>2) It is safe to walk after dark | 1 strongly disagree - 5 strongly agree |

* General health (SF-8) and quality of life (EQ-5D) were calculated as a weighted summary of multiple items, therefore, the range of this varied across settings. In our study, range for EQ-5D ranges from-0.594 to 1 as similar to UK's value set [8]. While there is no consensus in reported range, the general health (SF-8) of PCS and MCS in our study ranged from 11 to 72.

#### Table S2: Description of covariates

| Covariates | Description |
|---|---|
| **Gender** | 1 – Male, 0- Female |
| **Age** | Based on reported birth year, continuous from 16 onwards |
| **Marital status** | 1 – single, 2 – Married/ Cohabited, 3 – Other (separated, divorced, widowed) |
| **Education** | 1 – High level: obtained at least a degree or A-level<br>0 – Low level: Apprenticeship/ GCSE or lower |
| **Employment** | 1 – employed<br>0 – Unemployed/ economic inactive |
| **Obesity** | 1 – Normal (BMI<25)<br>2 – Overweight (BMI from 25-30)<br>3 – Obese (BMI>30) |
| **Smoking** | 1 – Current smoker<br>2 – Ex-smokers<br>3 – Never smoker |
| **Drinking** | 1 – Currently drinking<br>0 – Never/ very occasionally |
| **Long term illness** | 1 – Yes<br>0 – No |
| **Income** | 5 quintiles from 1 poorest to 5 richest. Each wave, participants were asked about their estimated gross income from a number of income brackets (which were inflation-adjusted each wave). For the convenience of data merge, the income variable was converted into 5 quintiles and merged across three waves. |
| **Distance to CCG** | The distance from respondents' address to the nearest accessible point of CCG infrastructure: 1) <400m; 2) 400 – 800m; 3) >800 – 1200m; 4) >1200m |
| **Area based deprivation** | Obtained using 2010 Northern Ireland Multiple Deprivation Measure [9]. Since the study sample was selected based on electoral wards, this led to the missing of a few deprivation levels in the control group – this variable was only presented in Table 1 for descriptive purpose and not used in the final analysis. |

#### Appendix 2: Descriptive statistics of study outcomes across three waves

Descriptive statistics of study outcomes at the intervention and control site were shown below.

**Table S3: Descriptive results of study outcomes across three waves**

| Year | Intervention (weighted) | | | Control (weighted) | | |
|---|---|---|---|---|---|---|
| | **Wave 1** | **Wave 2** | **Wave 3** | **Wave 1** | **Wave 2** | **Wave 3** |
| *N =* | *1037* | *968* | *944* | *168* | *246* | *258* |
| **Physical activity** | | | | | | |
| **Meet the PA recommendation** | 73.1% (1.4%) | 65.5% (1.5%) | 72.7% (1.4%) | 67.9% (3.7%) | 71.4% (2.9%) | 74.2% (2.7%) |
| **% Had active travel during a week** | 64.4% (1.6%) | 66.9% (1.5%) | 73.1% (1.4%) | 53.5% (4.0%) | 66.5% (3.1%) | 69.6% (2.9%) |
| **Total time for physical activities** | 845.53 (1369.21) | 675.06 (1073.80) | 676.28 (959.60) | 1059.99 (1421.30) | 701.34 (1061.70) | 883.56 (1339.17) |
| **Time of vigorous-intensity PA** | 149.14 (489.70) | 112.81 (387.38) | 118.40 (335.71) | 154.61 (443.64) | 110.00 (400.87) | 171.80 (520.38) |
| **Time of moderate-intensity PA [a]** | 547.26 (830.39) | 449.45 (675.49) | 439.48 (610.51) | 750.76 (1117.07) | 481.34 (631.40) | 539.95 (754.75) |
| **Time of active travel** | 352.74 (724.97) | 261.90 (582.20) | 258.55 (546.08) | 534.86 (1024.82) | 333.11 (614.24) | 309.80 (623.32) |
| **Health and wellbeing** | | | | | | |
| **WEMWBS [b]** | 50.76 (9.40) | 51.65 (9.66) | 50.06 (9.29) | 51.89 (8.59) | 51.34 (9.06) | 50.35 (9.18) |
| **SF-8: Physical (PCS)** | 30.56 (11.59) | 29.72 (10.64) | 31.31 (11.01) | 30.76 (10.82) | 28.98 (9.85) | 30.30 (10.88) |
| **SF-8: Mental (MCS)** | 32.68 (12.39) | 30.56 (12.03) | 34.26 (11.78) | 31.63 (11.11) | 30.73 (11.40) | 34.26 (12.33) |
| **EQ-VAS [c]** | 73.90 (21.46) | 64.03 (32.14) | 64.67 (31.83) | 76.71 (16.92) | 54.92 (38.00) | 59.30 (32.23) |
| **EQ5D3L** | 0.81 (0.28) | 0.83 (0.27) | 0.76 (0.28) | 0.83 (0.25) | 0.87 (0.21) | 0.77 (0.27) |
| **Social capital** | | | | | | |
| ***Local area trust [d]*** | 3.39 (0.64) | 3.54 (0.53) | 1.61 (0.68) | 3.56 (0.50) | 3.71 (0.43) | 1.36 (0.54) |
| ***Social network [e]*** | 3.99 (0.63) | 3.89 (0.65) | 2.96 (0.56) | 4.00 (0.57) | 3.96 (0.66) | 3.09 (0.55) |
| **Environmental perceptions** | | | | | | |
| ***Attractive [f]*** | 3.54 (0.87) | 3.85 (0.66) | 3.89 (0.72) | 3.80 (0.72) | 3.97 (0.52) | 4.02 (0.66) |
| ***Traffic [f]*** | 2.74 (0.77) | 2.92 (0.64) | 3.00 (0.72) | 2.89 (0.82) | 2.98 (0.61) | 3.06 (0.77) |
| ***Amenities [f]*** | 3.84 (0.56) | 3.87 (0.59) | 3.50 (0.47) | 3.67 (0.77) | 3.71 (0.69) | 3.35 (0.56) |
| ***Safety [f]*** | 3.50 (0.88) | 3.70 (0.75) | 2.15 (0.76) | 3.71 (0.78) | 3.86 (0.70) | 2.02 (0.76) |

**a**- Refers to moderate work & recreational activities and not including active travel (walking/ cycling);
**b**- WEMWBS scale from 14–70 with higher scores indicating better mental wellbeing;
**c** - EQ-VAS scale from 0–100 with higher scores indicating better health;
**d** - 1=very big problem to 4=not a problem at all; **e**- 1=never to 5=most days, **f**- 1=strongly disagree to 5=strongly agree.

#### Appendix 3: Population trends on physical activity level and mental wellbeing

The following trend is obtained from the latest result of the 2023/2024 Health Survey Northern Ireland (available at https://www.health-ni.gov.uk/publications/health-survey-northern-ireland-first-results-202324, accessed on 3 Nov 2025). For physical activity, data was not available on 2010/11 and 2017/18 so data from the closest timepoint was selected.

| % Meet the PA recommendation | 2012/13 | 2016/17 | 2023/24 | Significant difference | |
|---|---|---|---|---|---|
| | | | | 2012/13 & 2023/24 | 2016/17 & 2023/24 |
| **Overall** | 53.7% | 57.3% | 54.6% | ↔ | ↔ |
| **16-24** | 68.9% | 71.6% | - | - | - |
| **25-34** | 67.5% | 66.7% | 66.5% | ↔ | ↔ |
| **35-44** | 65.0% | 71.0% | 68.1% | ↔ | ↔ |
| **45-54** | 59.4% | 63.7% | 57.0% | ↔ | ↔ |
| **55-64** | 41.9% | 47.6% | 42.8% | ↔ | ↔ |
| **65-74** | 36.6% | 46.9% | 47.7% | ↑ | ↔ |
| **75+** | 11.6% | 15.8% | 26.3% | ↑ | ↑ |

| WEMWBS score | 2010/11 | 2017/2018 | 2023/24 | Significant difference | |
|---|---|---|---|---|---|
| | | | | 2010/11 & 2023/24 | 2018 /19 & 2023/24 |
| **Overall** | 49.71 | 51.58 | 50.74 | ↑ | ↓ |
| **16-24** | 50.05 | 51.62 | 51.21 | ↔ | ↔ |
| **25-34** | 49.56 | 52.91 | 51.13 | ↑ | ↔ |
| **35-44** | 49.01 | 51.51 | 50.64 | ↑ | ↓ |
| **45-54** | 48.38 | 50.78 | 49.83 | ↑ | ↓ |
| **55-64** | 49.54 | 50.12 | 49.64 | ↔ | ↔ |
| **65-74** | 51.77 | 52.52 | 52.29 | ↔ | ↔ |
| **75+** | 51.47 | 51.99 | 51.16 | ↔ | ↔ |

#### Appendix 4: Changes in study outcome over 6 years post-intervention

| | Wave 3 vs. wave 2 (post-intervention change) | | | | | |
|---|---|---|---|---|---|---|
| | INTERVENTION | | | CONTROL | | |
| | Mean difference* | 95%CI | P | Mean difference* | 95%CI | P |
| **Physical activity** | | | | | | |
| % Meet the PA target [a] | 1.19 | (0.90, 1.57) | 0.23 | 1.19 | (0.70, 2.01) | 0.52 |
| % Active travel [a] | 1.21 | (0.95, 1.55) | 0.12 | 1.53 | (0.87, 2.70) | 0.14 |
| Total PA time [b] | -83.3 | (-224.8, 58.2) | 0.25 | 166.3 | (-160.2, 492.8) | 0.32 |
| Time in vigorous-intensity PA | -19.1 | (-58.2, 19.9) | 0.34 | 55.8 | (-46.7, 158.3) | 0.29 |
| Time in moderate-intensity PA [c] | -28.3 | (-103.8, 47.3) | 0.46 | -53.3 | (-173.5, 66.8) | 0.38 |
| Active travel time | -12.6 | (-37.4, 12.1) | 0.32 | 109.5 | (52.3, 166.7) | <0.001 |
| **Health and wellbeing** | | | | | | |
| WEMWBS score [d] | -1.79 | (-2.66, -0.92) | <0.001 | -1.23 | (-2.76, 0.29) | 0.11 |
| SF-8: Physical component (PCS) | 2.04 | (1.05, 3.03) | <0.001 | 1.04 | (-0.33, 2.41) | 0.14 |
| SF-8: Mental component (MCS) | 3.48 | (2.45, 4.50) | <0.001 | 2.52 | (-0.41, 5.45) | 0.09 |
| EQ-VAS [e] | -0.46 | (-6.85, 5.92) | 0.89 | 2.51 | (-10.17, 15.18) | 0.70 |
| EQ5D-3L | -0.07 | (-0.10, -0.05) | <0.001 | -0.09 | (-0.13, -0.05) | <0.001 |
| **Social capital** | | | | | | |
| Local area trust [f] | -1.97 | (-2.12, -1.81) | <0.001 | -2.33 | (-2.52, -2.14) | <0.001 |
| Social network [g] | -0.95 | (-1.01, -0.89) | <0.001 | -0.87 | (-0.99, -0.75) | <0.001 |
| **Perceptions of environment** | | | | | | |
| Attractive [h] | -0.003 | (-0.09, 0.08) | 0.95 | 0.07 | (-0.05, 0.18) | 0.27 |
| Traffic [h] | 0.10 | (0.03, 0.16) | 0.004 | 0.07 | (-0.07, 0.20) | 0.35 |
| Amenities [h] | -0.41 | (-0.48, -0.35) | <0.001 | -0.32 | (-0.50, -0.15) | <0.001 |
| Safety [h] | -1.60 | (-1.78, -1.43) | <0.001 | -1.81 | (-2.07, -1.55) | <0.001 |

* Mean difference: % meeting the PA target and % had active travel reported ORs from the multilevel mixed-effect logistic model. For other outcomes, coefficients from multilevel mixed-effect linear regressions were reported.

a – reported having walking/ cycling during a typical week;

b - reported in equivalent of minutes of moderate-intensity PA per week;

c - Moderate activities at work + recreational activities, not including active travel;

d - WEMWBS scale from 14–70;

e - EQ-VAS scale from 0–100;

f - 1=very big problem to 4=not a problem at all;

g - 1=never to 5=most days;

h - 1=strongly disagree to 5=strongly agree.
